# A prospective observational cohort study to identify inflammatory biomarkers for the diagnosis and prognosis of patients with sepsis

**DOI:** 10.1101/2021.06.16.21259042

**Authors:** Valentino D’Onofrio, Dries Heylen, Murih Pusparum, Inge Grondman, Johan Vanwalleghem, Agnes Meersman, Reinoud Cartuyvels, Peter Messiaen, Leo AB Joosten, Mihai G. Netea, Dirk Valkenborg, Gökhan Ertaylan, Inge C. Gyssens

## Abstract

**Background:** Sepsis is a life-threatening organ dysfunction. A fast diagnosis is crucial for patient management. Proteins that are synthesized during the inflammatory response can be used as biomarkers, helping in a rapid clinical assessment or an early diagnosis of infection. The aim of this study was to identify biomarkers of inflammation for the diagnosis and prognosis of infection in patients with suspected sepsis.

**Methods:** In total 406 episodes were included in a prospective cohort study. Plasma was collected from all patients on the first day of a new episode. Samples were analysed using a 92-plex proteomic panel based on a proximity extension assay with oligonucleotide-labelled antibody probe pairs (OLink, Uppsala, Sweden). Supervised and unsupervised differential expression analyses and pathway enrichment analyses were performed.

**Results:** Supervised differential expression analysis revealed 21 proteins that were significantly lower in circulation of patients with viral infections compared to patients with bacterial infections. More strongly, higher expression levels were observed for 38 proteins in patients with high SOFA scores (>4), and for 21 proteins in patients with worse outcome. These proteins are mostly involved in pathways known to be activated early in the inflammatory response. Unsupervised, hierarchical clustering confirmed that inflammatory response was more strongly related to disease severity than to aetiology.

**Conclusion:** Several differentially expressed inflammatory proteins were identified that could be used as biomarkers for sepsis. These proteins are mostly related to disease severity. Within the setting of an emergency department, they could be used for outcome prediction, patient monitoring, and directing diagnostics.

**Registration:** clinicaltrial.gov identifier NCT03841162

## Introduction

Sepsis is a life-threatening organ dysfunction caused by a dysregulated host response to infection [1, 2]. Septic shock is a subset of sepsis in which profound circulatory, cellular, and metabolic abnormalities are associated with a greater risk of mortality than with sepsis alone [1]. In patients with sepsis, the fast initiation of antibiotic therapy is crucial and each hour delay results in increased risk of mortality [3]. The causative event is an invading pathogen, most frequently *Escherichia coli, Staphylococcus aureus, Streptococcus pneumoniae*, and *Klebsiella species*, but also influenza virus. Pneumonia, urinary tract infections, and intra-abdominal infections commonly result in sepsis, although the initial site of infection is not always known, e.g., in primary bloodstream infections (BSI) [2, 4]. However, infection sites are related to different causative pathogens and require different antimicrobial treatment.

Organ dysfunction resulting from infection is represented in the new definition of sepsis and defined by an increased sequential organ failure assessment (SOFA) score and is often caused by the dysregulated host response. An increase in SOFA score of 2 or more is associated with an in-hospital mortality of 10% [1]. The host response can be heterogeneous and is characterized not only by excessive inflammation, but also by immune suppression. Ultimately, the host response can be unbalanced and harmful leading to failure to return to homeostasis [5]. The increasing knowledge on host response in sepsis allows for the measurement of inflammation to identify blood biomarkers for a better diagnosis and prognosis of patients with various infections. Several biomarkers for inflammation and infection exist, although they mostly provide an indication on the clinical state of the patient while lacking sensitivity or specificity needed to be used as a diagnostic tool [6]. Two widely used markers are white blood cell count (WBC) and C-reactive protein (CRP). Both are positive inflammatory markers and indicate bacterial infection, although they lack specificity [7, 8]. Leukocytosis is associated with bacterial infection, and highly increased CRP levels correlate with a positive sensitivity for bacterial infection [6-9]. Additionally, procalcitonin (PCT), a pro-inflammatory biomarker released by monocytes and macrophages, correlates with inflammation intensity and highly increased concentrations are seen in bacterial infections [7-9]. Although PCT is able to discriminate between bacterial and viral community-acquired pneumonia, a multi-marker approach seems more reliable [6, 8]. Biomarkers can help in providing a rapid clinical assessment of patients and predict disease severity. Therefore, they could improve outcomes by rapidly guiding triage and the start of adequate treatment [9, 10]. A faster diagnosis of the infection and the likely causative pathogen, will lead to a faster start of targeted antimicrobial therapy, thereby reducing selective pressure for antimicrobial resistance (AMR). A faster prognostic assessment will guide closer patient monitoring. The aim of this study was to identify biomarkers of inflammation for determining the aetiology of the infection and its prognosis in patients with suspected sepsis.

## Materials and Methods

### Study design and study patients

This study was part of a prospective observational cohort study performed between February 2019 and April 2020 at a 981-bed teaching hospital (clinicaltrial.gov identifier NCT03841162).

Adult patients presenting with suspected sepsis at the emergency department (ED), the department of infectious diseases/nephrology, or the haemodialysis unit could participate in the study. All patients from whom blood cultures were drawn were considered to have suspected sepsis. Patients were included after collection of the first set of blood cultures. Patients could be included multiple times if they developed a new suspected sepsis episode. A new episode was defined as a minimal interval of seven days between positive cultures with the same pathogen or at least 24 hours between positive cultures with different organisms from the same site.

This is a sub-study in which patients with primary BSI, pneumonia, influenza, urosepsis, or other secondary BSI were included for biomarker identification between February 2019 and March 2020. An analysis on risk factors for patient outcomes of the complete study cohort (1690 episodes of suspected sepsis) was performed earlier [11]. The methods of patient inclusion, microbiological diagnostics, and definitions of infection diagnoses were identical in both analyses.

### Sample collection

Blood cultures, EDTA samples for complete blood count (CDC) and serum and heparin plasma samples for basic laboratory parameters assessing organ function were obtained for each patient at the start of each new episode. Two 9 mL EDTA blood samples were drawn from the same venepuncture. EDTA samples were stored at 4°C until written informed consent was obtained for a maximum of three days. Samples were transferred to the University Biobank Limburg (UBiLim) and centrifuged at 400g for 10 minutes. Plasma was separated and centrifuged at 1500g for 10 minutes. Four aliquots of 500μL plasma were stored at UBiLim [12] for each patient at −80°C until further analysis.

### Microbiological diagnostics

Blood cultures were performed for all patient episodes using the BACTEC FX (Becton Dickinson) system. Bacterial identification was done by MALDI-TOF Biotyper (Bruker). Susceptibility testing was done by the Phoenix system TM 100 (Becton Dickinson). Blood cultures were processed 24h/day, 7d/week. Other microbiological diagnostics were performed if deemed relevant by the treating physician. This included cultures of urine, lower respiratory tract and samples of specific foci, urinary antigen tests for *Streptococcus pneumoniae* and *Legionella pneumophila*, and PCR for respiratory pathogens on nasopharyngeal swabs.

### Data collection

Relevant data was extracted from patients’ electronic medical files at the start of each new episode. SOFA-score at the start of a new episode was calculated for all patients [1, 13]. Recorded patient outcomes were in-hospital mortality, intensive care unit (ICU) admission at any time during hospital admission, hospital and ICU length of stay (LOS), and the presence of bacteraemia. A composite parameter for worse outcome was made. Patients who were admitted to the ICU or who died during hospitalization were classified in the worse outcome group.

### Definitions

The final diagnosis of infection was extracted from the treating physicians’ discharge letter and structured and validated according to infectious diseases definitions [1, 14-17] by an experienced ID physician (I.C.G.) not involved in the care or consultations of study patients. Sepsis-3 definitions were used to define the presence of sepsis, i.e., all patients with a SOFA score of ≥2, and septic shock at the start of an episode [1]. Severe disease was defined as SOFA score of >4 and compared with less severe disease (SOFA <2). For all infection diagnoses, patients with a SOFA score of one or higher were included in this analysis. Positive blood cultures were classified as true bacteraemia or as contamination according to CDC guidelines [14]. Positive blood cultures with skin flora, such as coagulase-negative staphylococci, were considered as contaminated when less than two blood cultures bottles from one patient were positive for skin flora. Patients with a positive blood culture with these organisms and a clinical suspicion of an infection of central venous catheters or surgically implanted prosthetic material were considered to have true bacteraemia. Primary BSI was defined as true bacteraemia, without a focus of infection. Patients with a central-line associated BSI (CLABSI) or with confirmed endocarditis were included in this group. Pneumonia was defined as an acute symptomatic infection of the lower respiratory tract whereby a new infiltrate is demonstrated [15]. Influenza was defined by a positive influenza PCR test. Patients with both influenza and pneumonia were classified as having influenza and were further classified as influenza with or without pneumonia based on the presence of chest X-ray abnormalities. Other viral causes of pneumonia were not detected in this population. Therefore, pneumonia without the presence of influenza was classified as bacterial pneumonia. Secondary bacteraemia patients had true bacteraemia with a urinary tract focus [16], an intra-abdominal focus or a skin and skin structure infection.

### Protein identification

EDTA plasma from patients was analysed using a proteomic multiplex assay (Olink, Uppsala, Sweden), which is a proximity extension assay with oligonucleotide-labelled antibody probe pairs [18]. Samples were analysed with the inflammation panel, which enables analysis of 92 inflammation-related proteins. Briefly, a pair of oligonucleotide-conjugated antibodies to each protein is added to 50μL EDTA plasma. Both antibodies are in close proximity when an antibody-protein-antibody sandwich is formed. This allows hybridization of the oligonucleotides, and an extension reaction forms a unique sequence. The sequences are quantified by qPCR.

### Statistical analyses

Descriptive statistics were used to analyse patient’s characteristics. Continuous data are shown as median (interquartile range (IQR)). Categorical data are reported as number and proportion. A p-value of <.05 was considered statistically significant. Data on protein levels were analysed as normalised protein expression (NPX on a log2 scale). Normalization across batches was performed using bridging samples according to manufacturer’s instructions. Supervised analyses using Welch’s t-test were done to search for differences according to aetiology (influenza vs. bacterial infection), severity (SOFA score >4 versus SOFA score <2) and outcome (worse versus less severe outcome). This was followed by unsupervised hierarchical clustering and pathway enrichment analyses. Elastic net regression was performed to search for the best biomarker predictors of different outcomes. All analyses were performed using R. Statistical methods are detailed in the additional file.

## Results

### Patient Characteristics

In total, 406 episodes of suspected sepsis in 398 patients were included. Table 1 shows the characteristics of the cohort. The median age was 74 years, and 59.9% patients were male. Median Charlson Comorbidity Index (CCI) was 2 (2-9) with hypertension (27.1%), chronic kidney disease (26.8%), and cardiac comorbidities (23.4%) being the most frequent. Primary BSI was present in 17.7% of episodes, secondary bacteraemia including urosepsis in 31.3%, pneumonia in 31%, and influenza in 20%. Median SOFA score at the start of an episode was 2 (2-4). Septic shock was present in 1% of episodes and bacteraemia in 53.9%. Median LOS was 7 (4-13) days, and there was an ICU admission rate of 17.7% with a median ICU LOS of 4 (2-9) days. In-hospital mortality was 10.8%. Therefore, 96 (23.6%) patients were classified in the worse outcome group and 310 (76.4%) patients in the less severe outcome group.

**Table 1.** Patient demographics and characteristics, diagnosis of infection, disease severity and patient outcomes

|  | **Total**<br>**n = 406 episodes** |
| --- | --- |
| **Demographics** |  |
| Age (years, median (IQR)) | 74 (64-83) |
| Gender (male) | 243 (59.9) |
| Department of inclusion |  |
| Emergency department | 381 (93.8) |
| Infectious diseases | 16 (3.9) |
| Haemodialysis | 9 (2.2) |
| **Comorbidities** |  |
| CCI (median (IQR)) | 2 (0-3) |
| Cardiac | 95 (23.4) |
| Hypertension | 110 (27.1) |
| Cerebrovascular disease | 48 (11.8) |
| Chronic pulmonary disease | 65 (16.0) |
| Chronic kidney disease | 109 (26.8) |
| Liver disease | 18 (4.4) |
| Dementia | 28 (6.9) |
| Diabetes | 82 (20.2) |
| Solid malignancies | 84 (20.7) |
| Haematological malignancies | 16 (3.9) |
| **Diagnosis of infection** |  |
| Primary bacteraemia | 72 (17.7) |
| Secondary bacteraemia | 127 (31.3) |
| Bacterial pneumonia | 126 (31.0) |
| Influenza | 81 (20.0) |
| **Disease severity\*** |  |
| SOFA (median, IQR) | 2 (2-4) |
| SOFA (1) | 73 (18.0) |
| SOFA ($\geq 2$) | 333 (82.0) |
| Septic Shock | 4 (1.0) |
| Bacteraemia | 217 (53.4) |
| **Outcome** |  |
| Length of stay (days, median (IQR)) | 7 (4-13) |
| ICU admission | 72 (17.7) |
| ICU length of stay (days, median (IQR)) | 4 (2-9) |
| Mortality | 44 (10.8) |
| Worse outcome<sup>#</sup> | 96 (23.6) |

\* Disease severity based on SOFA score at the start of a new episode. # Patients who were admitted to the ICU or who died during hospitalization were classified in the worse outcome group. All variables are presented as number (%) unless otherwise specified.

### Supervised differential expression analyses

Figure 1 shows a Venn diagram of all proteins that were significantly differentially expressed for three different groupings. In total, 28/92 proteins were significantly differently expressed when comparing patients with influenza (with or without pneumonia) with patients with bacterial infections. All but six of these proteins (CCL11, CXCL11, IFNγ, MCP-2, SCF, and TRAIL) were significantly less expressed in patients with influenza. Thirty-eight proteins were significantly differently expressed comparing patients with high SOFA score (>4) and patients with low SOFA score (<2). All but two proteins (AXIN1 and CXCL5) were significantly more expressed in patients with high SOFA score (>4). Additionally, 21/92 proteins were significantly different in patients with worse outcome compared to patients with less severe outcome. All proteins, except SCF, were significantly more expressed in patients with worse outcome. Nineteen of these proteins were also differentially expressed comparing patients with high and low SOFA score. Mean expression and p-values for the 3 groupings are shown in Additional files Table 2, 3, and 4 respectively.

**Figure 1.**
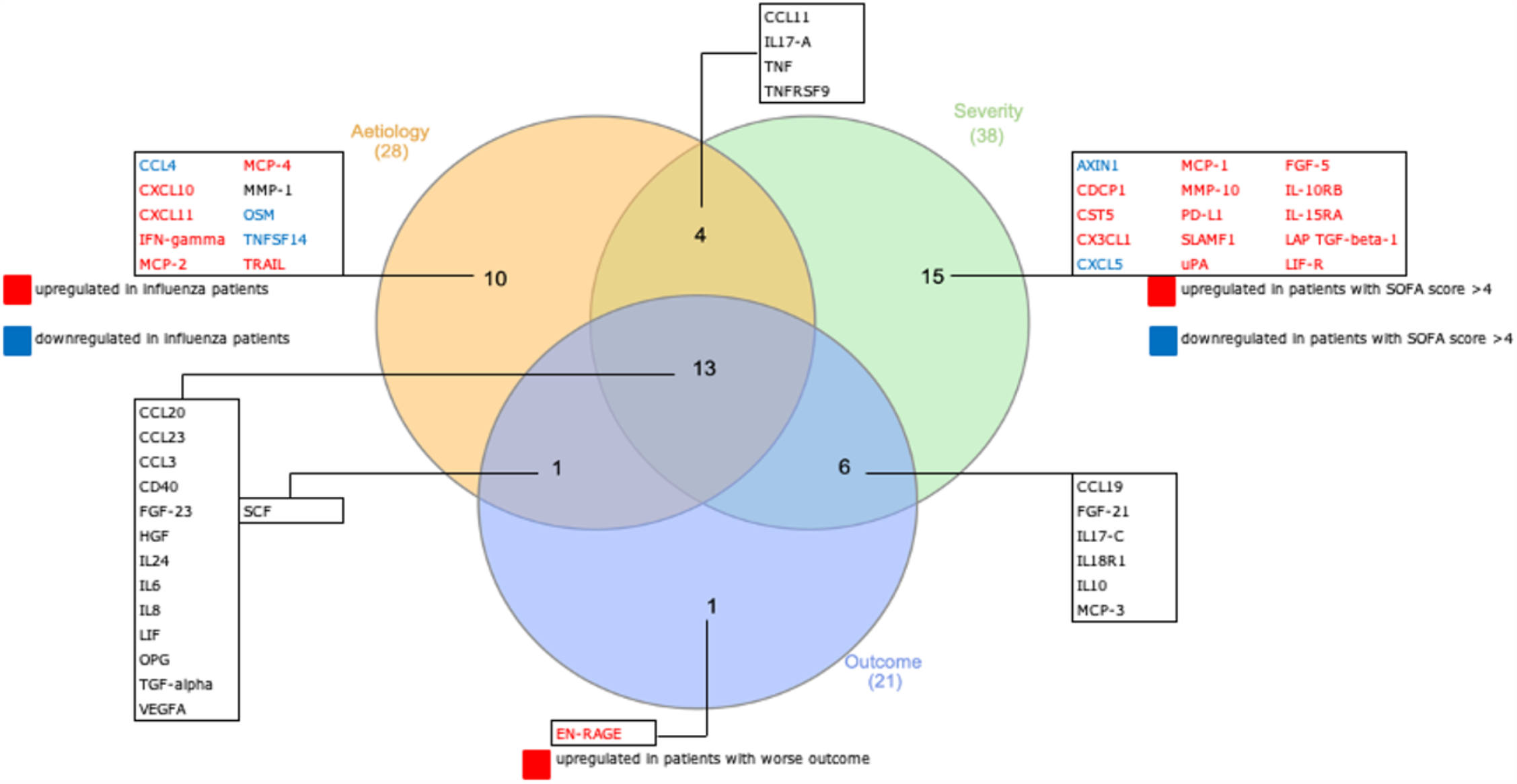
Venn diagram showing all differentially expressed proteins between three different groupings, together with overlapping findings. Three different groupings are: aetiology (influenza versus bacterial), severity (SOFA score <2 versus >4), and outcome (less severe outcome versus worse outcome).

### Unsupervised clustering

#### Principal component analysis

The first four main principal components explained 42% of the variance in the dataset (Additional files Table 1). Therefore, the following analyses were performed based on plotting these four components with each other. Gender, age, or batch confounding did not seem to be present in the normalized dataset (Additional files Figure 1).

Figure 2 shows the results of the principal component analyses. In order to look for distinct diagnosis-based clusters, inflammatory protein related profiles were initially checked by classifying patients in three diagnosis groups: influenza, bacterial pneumonia and other bacterial infection (Figure 2a). Bacterial pneumonia and influenza separated most strongly from each other as well as from other bacterial infections based on the PC2 axis. This indicates that the infection site or aetiology of infection can at least to some extent determine the inflammation associated protein expression in response to an infection. However, separation clearly experienced some overlap between the different diagnosis groups and was not absolutely distinct.

**Figure 2.**
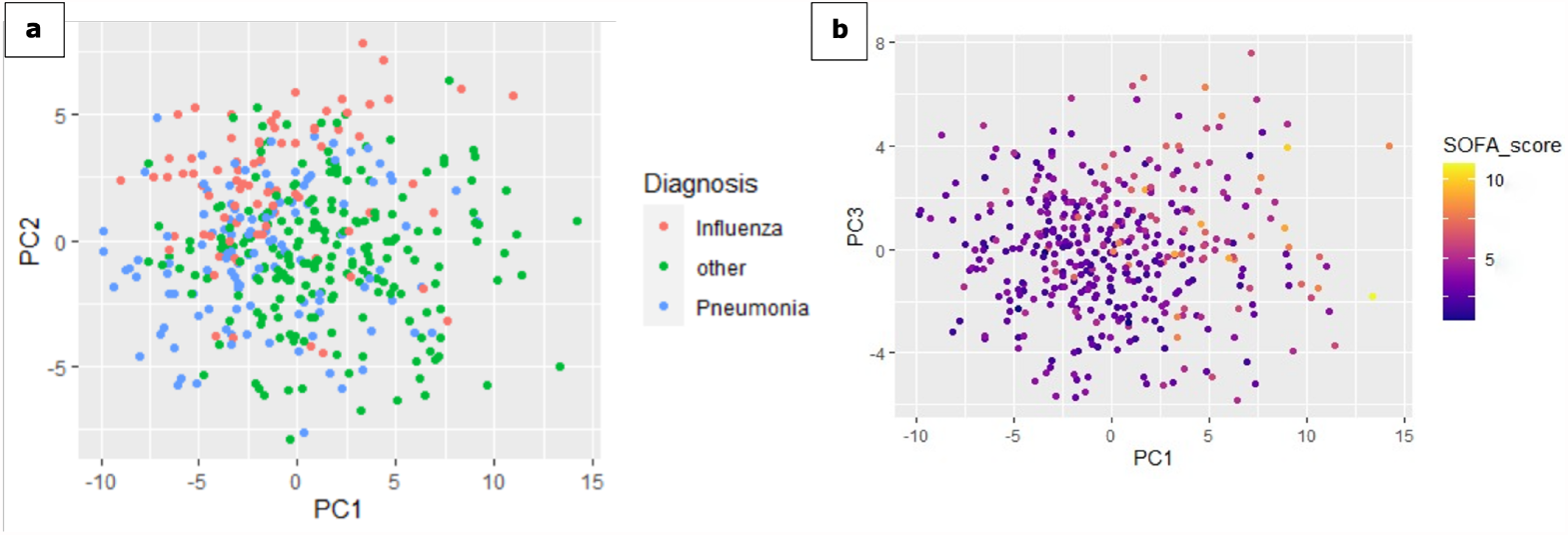
a: principal component analysis to detect clustering between patients with influenza (pink), bacterial pneumonia (blue) and other bacterial infections (green). b-c: principal component analysis to detect clustering based on SOFA score.

Interestingly, SOFA score seemed to grasp and identify the differences in protein expression accurately (Figure 2b). Patients with high SOFA scores clearly clustered together and differed from patients with low SOFA scores.

#### Hierarchical clustering

Results are shown in Figure 3. Hierarchical clustering confirmed that no confounding was induced by sex, age, or batch in the normalized dataset. Two distinct ‘Influenza’ groups could be distinguished at the extreme left and right of the cluster. The group on the left consisted mostly of patients with influenza without chest X-ray abnormalities, while the group on the right were mostly patients with influenza and chest X-ray abnormalities. Although at first it seemed that influenza clustered differently from bacterial pneumonia and other bacterial infections, thorough comparison of patients in different clusters revealed a pattern of severity that was considered as the underlying basis upon which the hierarchical clustering provided the presented dendrogram. In that same light, most patients with pneumonia and patients with other infections that clustered at the left of the dendrogram had lower SOFA scores than their variants situated at the right of the dendrogram. Furthermore, highest SOFA scores, and most BSIs (considered invasive disease) were seen in patients that clustered in the centre of the dendrogram. Additionally, when comparing outcomes, those patients with worse outcome mostly clustered in the middle, while the left and right clusters on the dendrogram mostly contained patients with less severe outcome. Therefore, hierarchical clustering could confirm supervised differential expression i.e., that some differences between viral (influenza) and bacterial infections were found, but that inflammatory proteins more strongly indicated disease severity.

**Figure 3.**
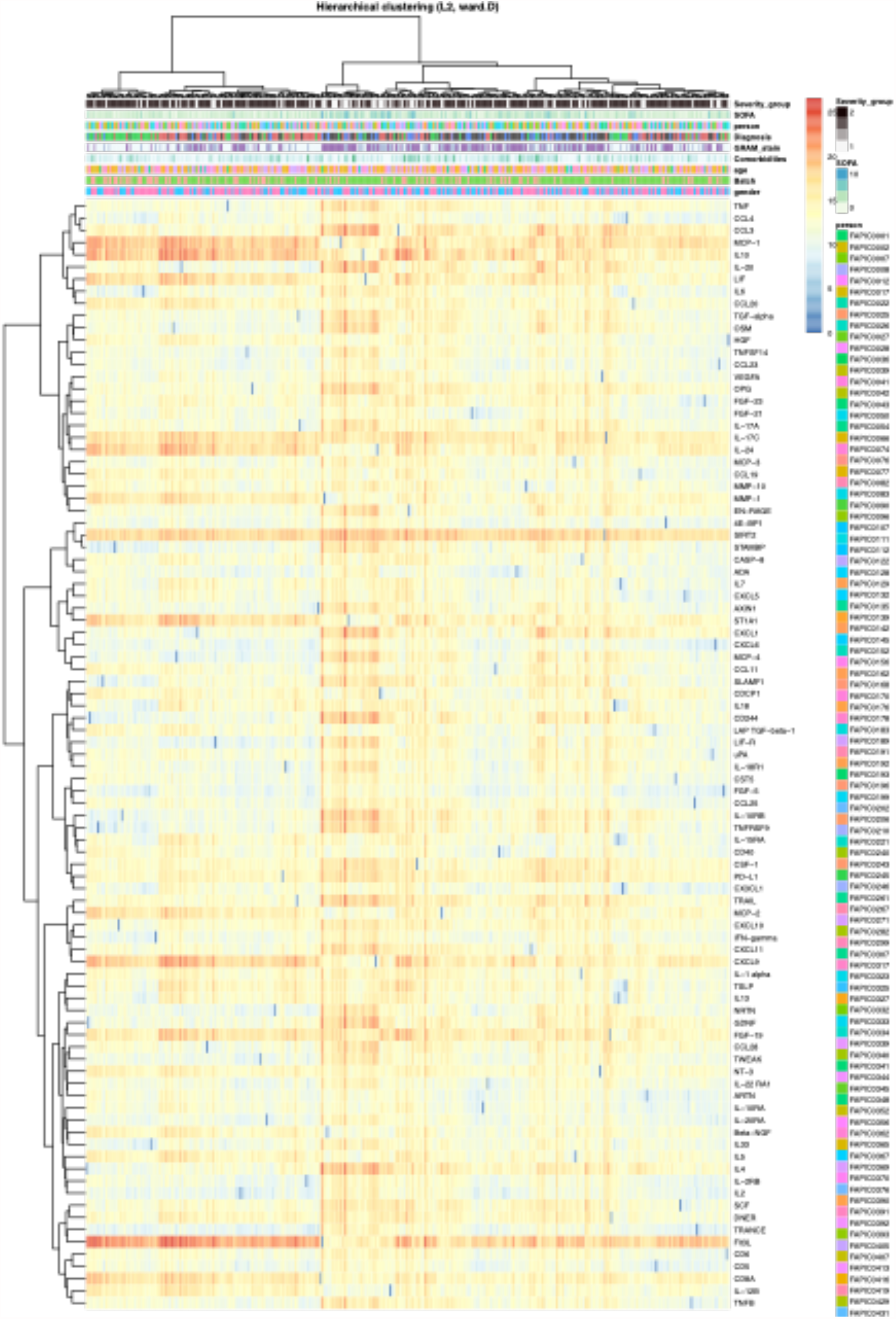
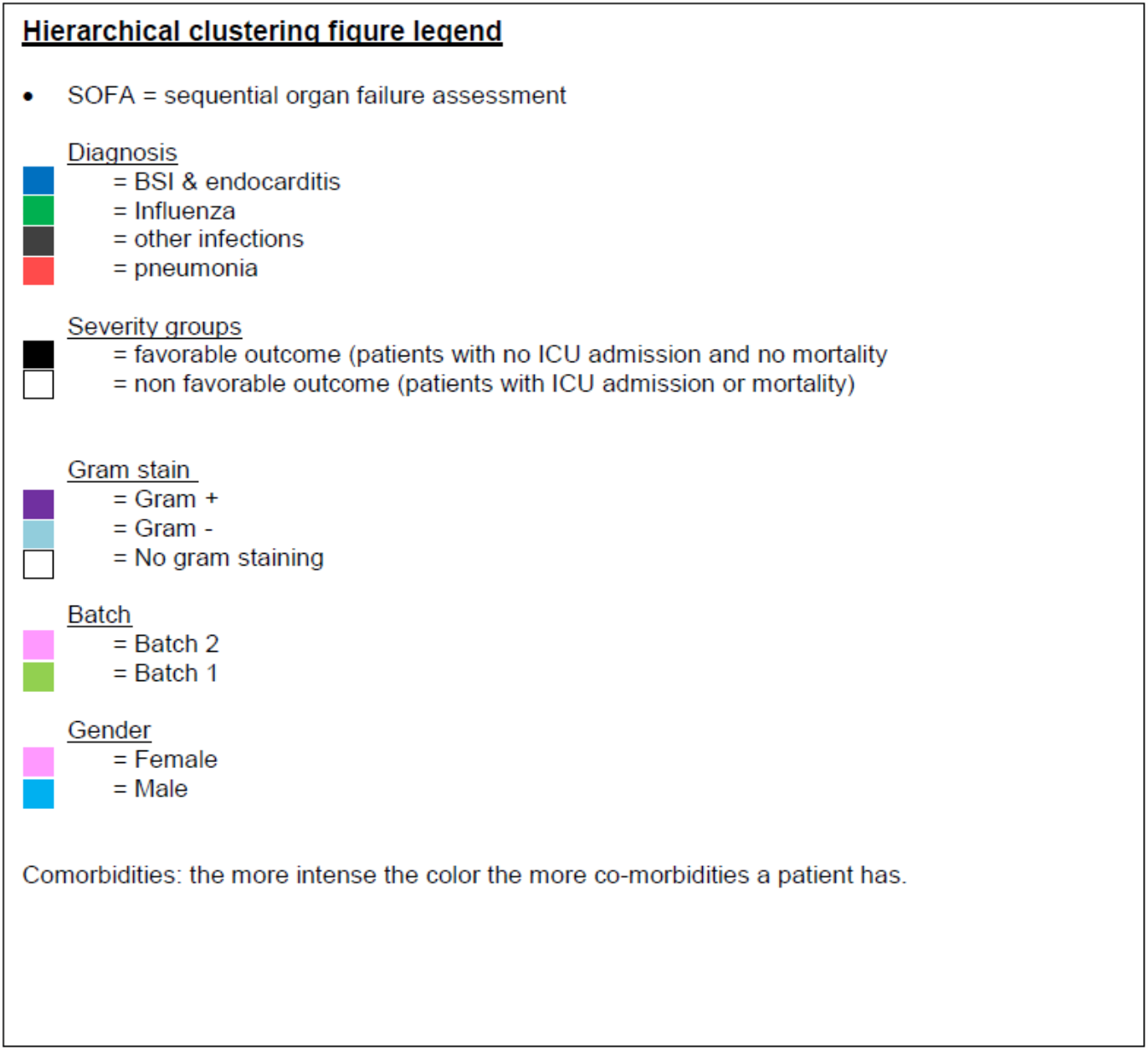
Hierarchical clustering plot.

### Pathway enrichment analysis

Pathway enrichment analysis was done to illustrate the molecular mechanisms involved in this population with sepsis as the inflammatory panel used has been compiled based on knowledge from a range of inflammatory diseases [19]. The most important pathway involving proteins differentially expressed between patients with influenza and patients with bacterial infections was HMGB1/TLR signalling pathway. In proteins differentially expressed between patients with worse and less severe outcome, ERBB family/HGF signalling and chemotaxis/CCR1 signalling were considered as the two most important pathways involved. These pathways show shared entities and provide a clear link with IL-10 signalling, IL-1 signalling, and HSP60 and HSP70/TLR signalling pathways, known to be involved in sepsis. In the latter three pathways, proteins were downregulated in patients with influenza and upregulated in patients with worse outcome, illustrating differences in disease severity. Figure 4 shows up-and downregulated proteins in the HSP60 and HSP70/TLR signalling pathway.

**Figure 4.**
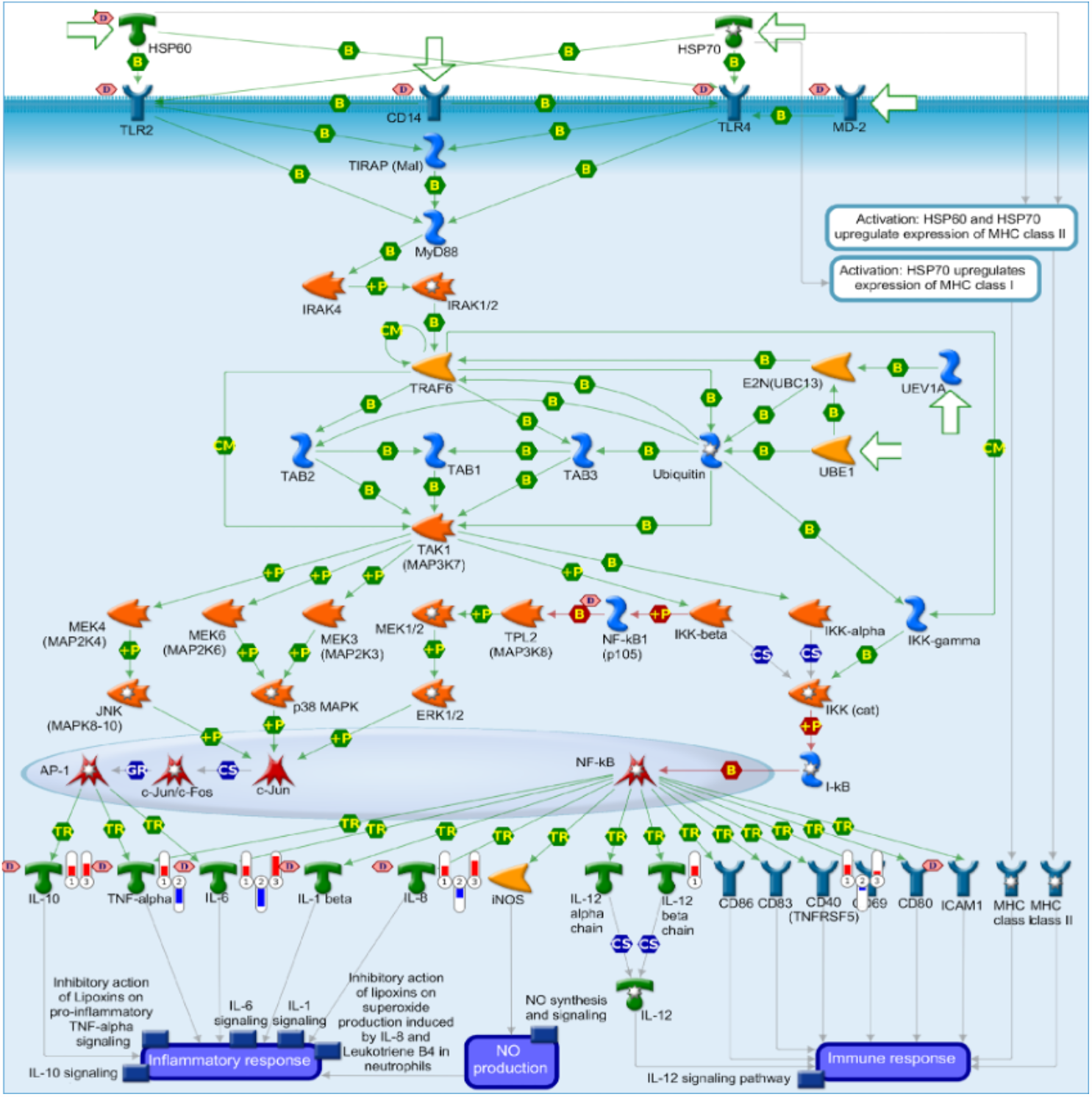
HSP60 and HSP70/TLR signalling pathway involved in the activation of the inflammatory response. Differentially expressed proteins in this population are specified. Legend: 1: present in inflammation panel but no significantly different expression 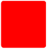 2: Differentially expressed protein upregulated in influenza patients, 3: Differentially expressed protein upregulated in patients with worse outcome 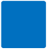 2: Differentially expressed protein downregulated in influenza patients, 3: Differentially expressed protein downregulated in patients with worse outcome Fold change in expression indicated by the ‘thermometer’, with the level of expression indicated by the height of the thermometer.

### Prediction Biomarkers

The accuracy of all elastic net models was tracked from the maximum number of biomarkers using a stepwise backward selection and is shown in Additional files Figure 2. Proteins in the most optimal model are shown in Figure 5. Six proteins could predict viral vs bacterial infection with an accuracy of 81%, six proteins could predict SOFA score >4 vs SOFA score <2 with an accuracy of 80%, and nine proteins could predict less severe outcomes from worse outcome with an accuracy of 75%.

**Figure 5.**
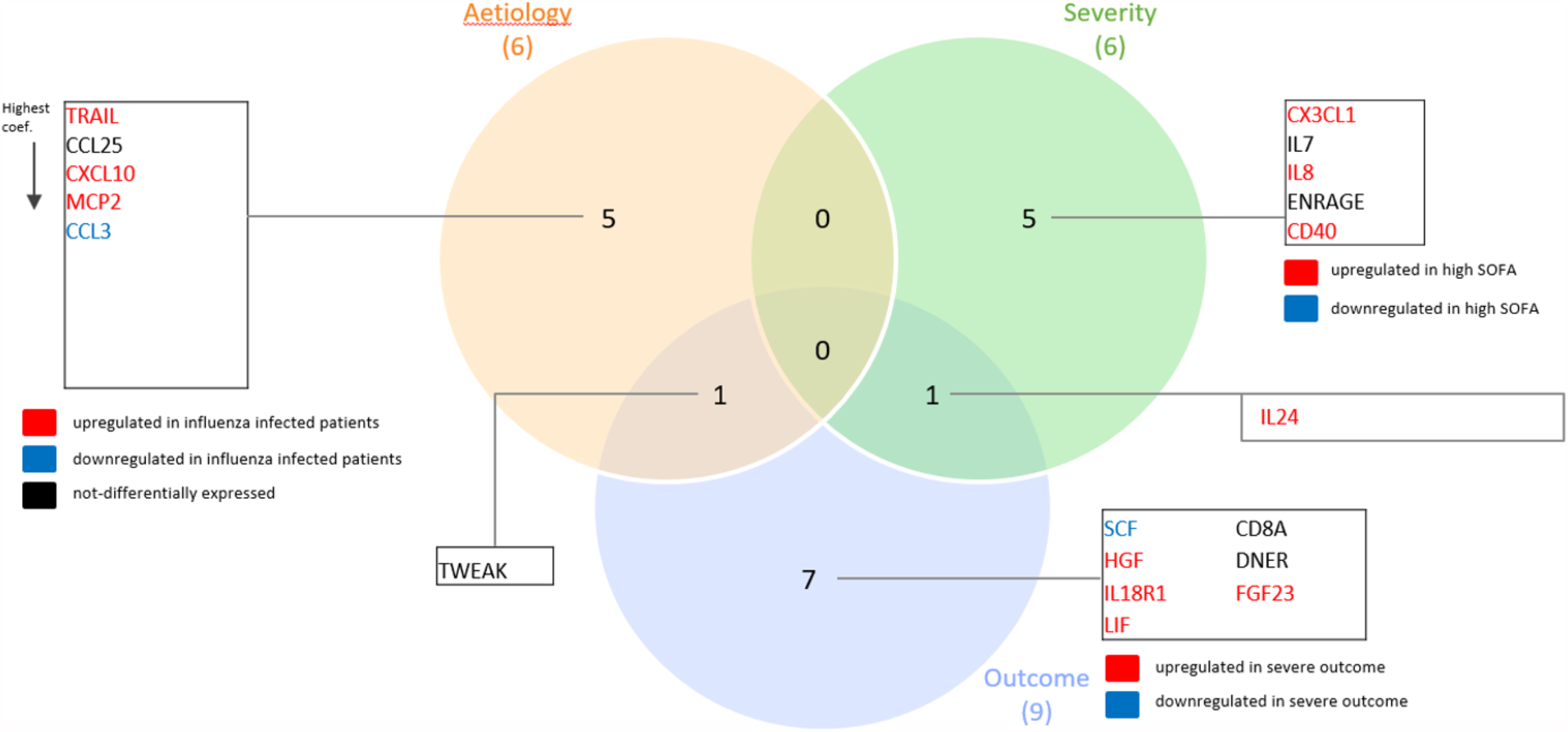
Proteins in the most optimal models, accurately predicting differences in aetiology, disease severity and outcome.

## Discussion

In this study of plasma biomarkers for early aetiologic diagnosis and prognosis of patients with suspected sepsis, one-third of a set 92 plasma proteins of inflammation were differentially expressed in viral infections (influenza) compared to infections of bacterial origin. However, patients with severe disease (high SOFA score) and worse outcome (mortality or ICU admission) had significantly higher protein concentrations compared to patients with less severe disease and less severe outcome. Furthermore, most identified proteins of the worse outcome group overlapped with those in patients with severe disease. This indicates that disease severity and patient outcome are related, and that these are associated with the inflammatory response to sepsis. Three models consisting of several protein biomarkers were built. They could accurately predict viral infection, severe disease and worse outcome. The aetiology model is particularly helpful for decisions whether to start empirical antibiotics in patients with suspected sepsis, the other two can point to the need for more intensive monitoring.

Extensive research has previously been performed to search for accurate and sensitive biomarkers for sepsis [20-22]. Most biomarkers have their limitations, especially in differentiating between sepsis and other inflammatory syndromes [20, 23]. Similar to our findings, plasma biomarkers are often shown to be more strongly related to disease severity rather than to the microbial aetiology of infection, and the majority of biomarkers have been evaluated for their use in the prognosis of patients with sepsis [20-22].

The availability of a commercial 92-plex panel for the detection of inflammation proteins allowed us to identify up to 38 differentially expressed proteins in this suspected sepsis population. Our results are in line with previous studies showing that a combination of several proteins more accurately differentiate between various disease severity profiles in sepsis and other infections, compared to single biomarkers [20, 24, 25]. However, our research, combining multiple biomarkers led to optimal models with as few biomarkers as possible for future applications in clinical practice.

Pathway enrichment analysis illustrated involvement of several pathways of early inflammatory response with several links to other pathways known to be involved in sepsis. In differentially expressed proteins, pathway enrichment analysis confirmed the involvement of RBB family/HGF signalling and chemotaxis/CCR1 signalling pathways for less severe or worse outcome and HMGB1/TLR signalling pathway for aetiology. These pathways are involved in inflammation, neutrophil recruitment and thus in the early immune response [26]. As a result, these pathways have similarities and shared entities with other pathways known to be involved in sepsis. For example, the IL-10 signalling pathway is involved in the inhibition of the immune response, and the IL-1 and HSP60 and HSP70/TLR signalling pathways are involved in activation of the innate and adaptive immune response. The observation was made that proteins in both inflammatory activation and inhibition are upregulated in patients with worse outcome, which is proof of the dysregulated immune response. An increase in early proteins and especially chemokines is also reported by others [27-30], suggesting that higher levels of chemokines are necessary because of reduced neutrophil migration velocity, eventually leading to an impaired immune response which is associated with disease severity and lower survival rates [28-30]. The identified pathways have also been shown to be related to disease severity in COVID-19 patients [27].

Indeed, the observation that inflammatory protein expression profile is related to sepsis severity, has recently been described in patients with COVID-19, using the same targeted proteomic approach [31]. Differentially expressed proteins in COVID-19 patients were involved in the same pathways found in the present study [24, 27, 31]. This strengthens our finding that these proteins could be more useful for the prognosis of patients with severe infections, rather than for the aetiologic diagnosis.

One major strength of this study is the use of a large cohort of patients who were prospectively included and uniformly defined. The results of this study show the importance of the host response in predicting patient outcome. The unicentric nature of this study is a limitation resulting in a difficult extrapolation to other centres. Furthermore, we did not compare our findings with a non-sepsis control population, and therefore lack information on basic expression levels. Last, there were no follow-up plasma samples during patients’ hospitalization, and we were not able to assess the impact of later events on patient outcome.

In conclusion, the inflammatory response of patients with viral infection differed from the response of those with bacterial infection. However, the inflammatory response correlated more strongly with disease severity and worse patient outcome. The differentially expressed inflammatory proteins play an important role in the early host response to infection. An increase in these proteins was seen for patients with worse outcome, indicating that an overactivated response is already present in early stages of infection. Several proteins could accurately predict influenza, disease severity, or worse outcome. Although the diagnostic potential of these biomarkers for distinguishing bacterial from viral infections is limited, their value for prognosis can be high. In the future, the differentially expressed proteins found in this study could be incorporated in a point of care test. Routine use of such a test at the ED could help in early outcome prediction, guide close monitoring, and direct diagnostics, ultimately leading to better management of patients with suspected sepsis.

## Data Availability

The datasets used and analyzed during the current study are available from the corresponding author on reasonable request. The data are not publicly available due to them containing information that could compromise research participant privacy/consent.

## Declarations

### Ethics approval and consent to participate

All procedures performed in studies involving human participants were in accordance with the ethical standards of the institutional and/or national research committee and with the 1964 Helsinki Declaration and its later amendments or comparable ethical standards. Documented approval for this study was obtained from the Ethics committees of Hasselt University and Jessa Hospital [18.106/infect18.03 and 19.51/infect.19.02]. Written informed consent was obtained from all participants.

### Consent for publication

Not applicable.

### Author’s contribution

VD designed, coordinated and executed the clinical cohort study and patient inclusion, did experimental work, collection and analysis of the data, and wrote the article. DH and MP analysed the data. IG did laboratory work. IG did laboratory and experimental work. JV, AM and RC collaborated in the clinical cohort study on the nephrology department, ED and microbiology department respectively, PM, LJ, and MN designed the study and did critical analysis of the manuscript. DV and GE designed the study analysis plan, did analysis of the data and did critical analysis of the manuscript. ICG designed and coordinated the study and analysis and did critical analysis of the manuscript. All authors read and approved the final manuscript.

## Acknowledgments

The human biological material used in this publication was stored in and released from the University Biobank Limburg (UBiLim) [12].

This study is part of the Limburg Clinical Research Center (LCRC) UHasselt-ZOL-Jessa, supported by the foundation Limburg Sterk Merk (LSM), Hasselt University, Ziekenhuis Oost-Limburg and Jessa Hospital.

This research has been presented as an e-poster during the European Conference of Clinical Microbiology and Infectious Diseases (ECCMID), July 2021

