## Supplementary materials for "A prospective observational cohort study to identify inflammatory biomarkers for the diagnosis and prognosis of patients with sepsis"

**Supplementary information**

**Supplementary Methods**

*Supervised differential expression analyses*

Differential expression analyses were performed with a Welch’s t test also known as the unequal variances t-test. Differences were searched for all 92 proteins across patients with influenza (with and without pneumonia) and patients with other bacterial infections. In addition, differences were analysed according to severity (high SOFA score >4 versus low SOFA score <2) and to outcome (worse versus less severe outcome). Pairwise comparisons for each protein were made according to Petrera *et al.* (1). The basic ‘stats’ R-package was used. A Bonferroni multiple testing correction was performed. A p-value < 0.05 was considered significantly differentially expressed.

*Unsupervised clustering*

Initial data exploration and data dimensionality reduction was done using principal component analysis (PCA). PCA is well known to highlight the most important aspects of data variability and de-emphasizes the others. The R-packages ‘ggplot2’, ‘ggreppel’ and ‘GGally’ were used to reveal the internal data structure by using an eigenvector-based multivariate approach.

Following supervised analysis, Hierarchical clustering, a commonly used unsupervised analysis method to check similarity between subjects, was used. In this analysis, samples that grouped together were considered to be more similar than samples from other patients in the cohort, i.e., they have comparable inflammatory protein patterns. Hierarchical clustering was performed using a Euclidean distance calculation method in combination with a Ward.D clustering method. R-packages ‘dplyr’, ,’randomcoloR’ and ‘ggplot2’ were loaded to complete the clustering. Clustering and distance calculation was performed by Ward.D clustering using Euclidean distance due to its ability to capture larger variations within the data avoiding splintered clustering. As the method is very sensitive to outliers, the control samples were excluded from the analysis.

*Pathway enrichment analysis*

Pathway enrichment analyses were performed on significantly differentially expressed proteins in MetaCoreTM. Protein Uniprot ID’s were searched in the light of their molecular function as well as their biological process. These analyses help gain mechanistic insight into gene lists and identify biological pathways that are enriched in a protein list, more enriched than would be expected by chance. Afterwards, proteins shared between these pathways and other pathways previously known to be involved in sepsis were integrated in the latter to allow for a better understanding of the role of these inflammatory biomarkers.

*Elastic Net*

To search for the best biomarker predictors of different outcomes, elastic net regression was selected as an alternative regularization technique that combines the L1 and L2-penalizations. This method does automatic variable selection and continuous shrinkage and produces a sparse model with better prediction accuracy, particularly in the two-class classification method (2).

Three categorical variables: aetiology (viral vs. bacterial infection) disease severity (based on SOFA score) and outcome (worse vs less severe) were proposed as vector of dependent variable $\boldsymbol{y}$ in three different models. The tuning parameter $\lambda$ controls the penalty factor and was chosen by 3-repeated 5-fold cross-validation. The models were trained in 80% of the data and the remaining was used for calculating the prediction accuracy and the area under the curve (AUC). All proteins were inserted in the starting model. The most optimal model was chosen based on the AUC. All estimations were performed in R *glmnet* and *ROCR* packages (3, 4).

**Supplementary Figures**

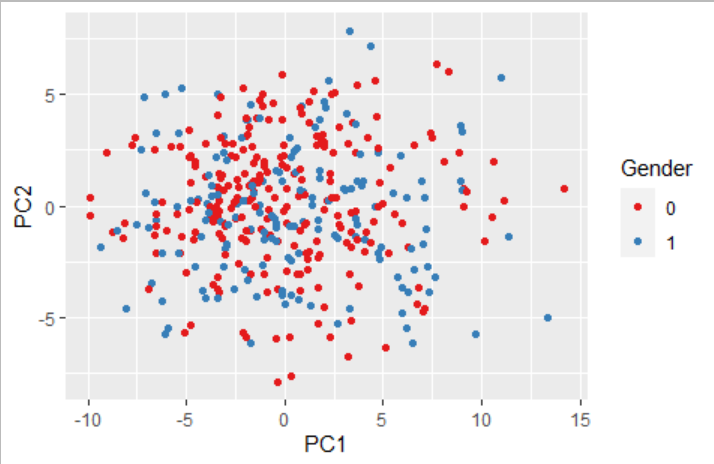

**a**

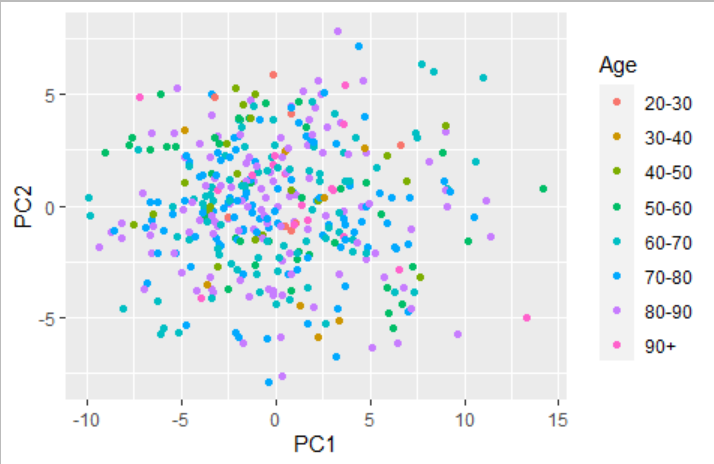

**b**

**c**

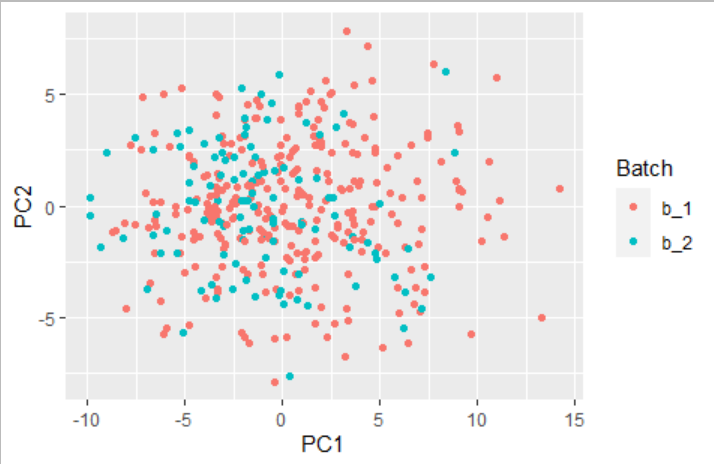

**Supplementary Figure 1**. Principal component analysis with groups based on gender (a), age (b), and batches (c). These were identified as not confounding.

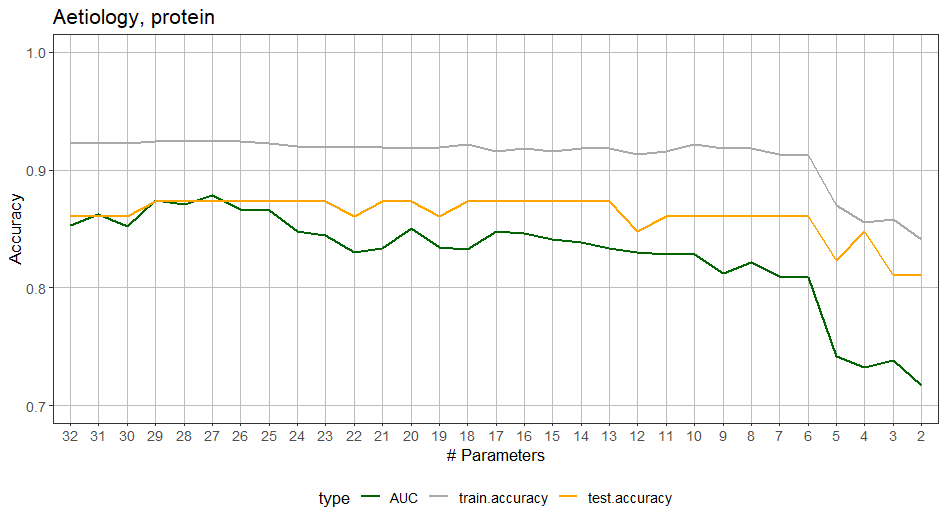

0.88

0.74

0.81

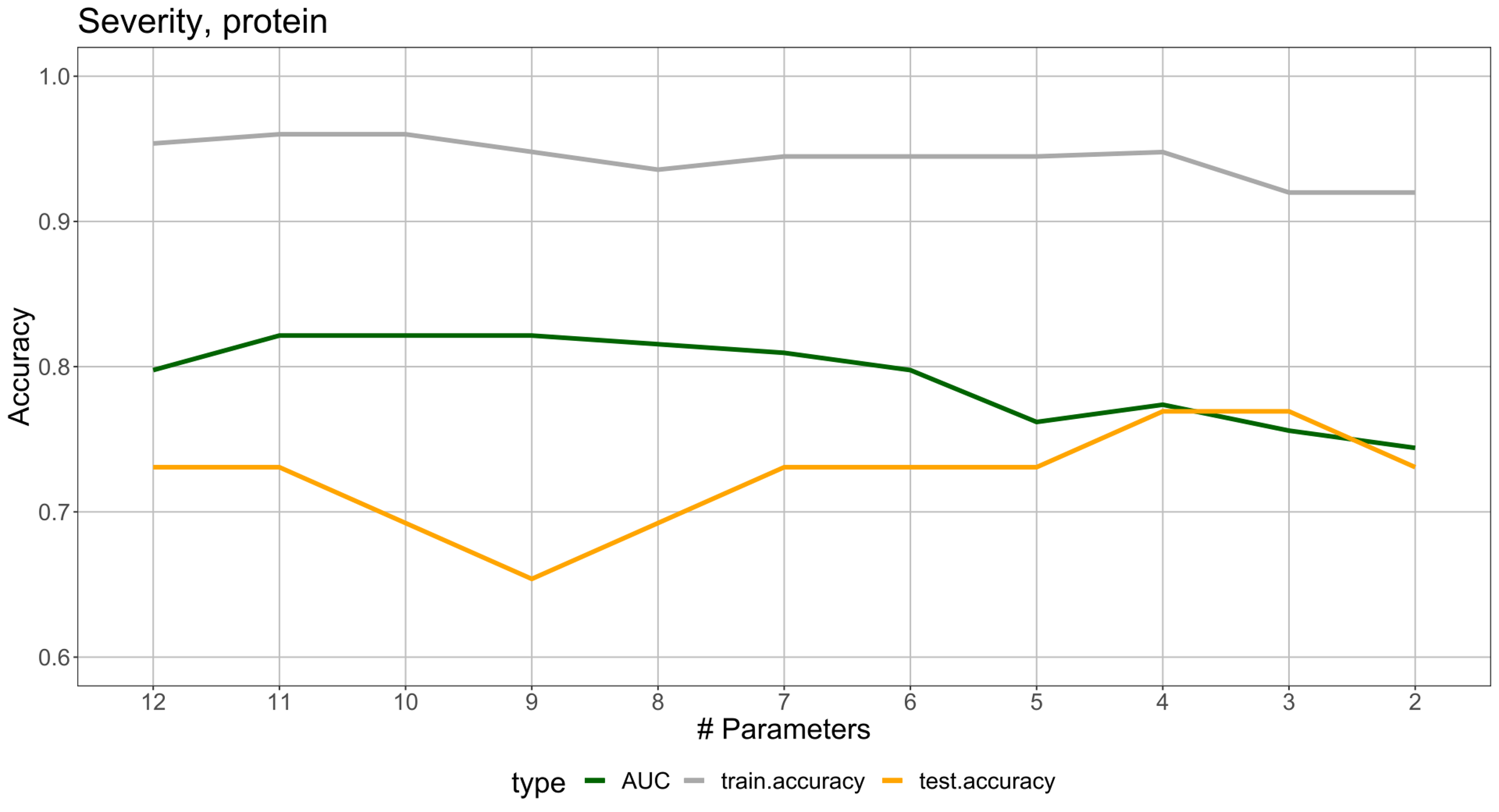

0.77

0.80

0.82

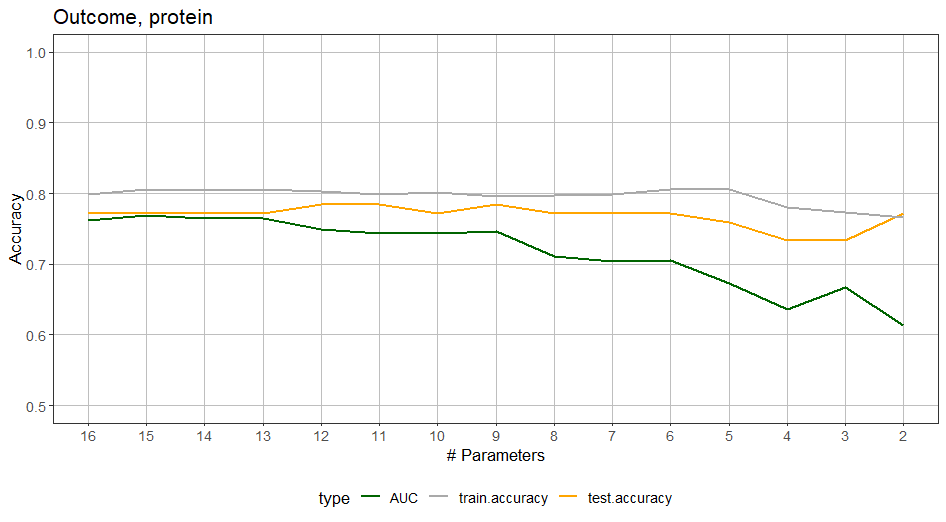

0.75

0.67

0.76

**Supplementary Figure 2.** Model optimization for aetiology, disease severity and outcome, starting from the maximum number of parameters resulting from the elastic net regression models. The most optimal model was chosen based on the highest area under the ROC curve (AUC) (approximately 80%) with the lowest number of proteins in the model. Aetiology: viral vs. bacterial sepsis, the most optimal model had an AUC of 81% with six proteins in the model. Disease severity: SOFA score >4 vs. SOFA score <2, the most optimal model had an AUC of 80% with six proteins in the model. Outcome: worse vs. less severe outcome, the most optimal model had an AUC of 75% with nine proteins in the model.

**Supplementary Tables**

**Supplementary Table 1.** The first five main principal components (PC). The first four PC explained 42% of observed variance.

|  | **PC1** | **PC2** | **PC3** | **PC4** | **PC5** |
| --- | --- | --- | --- | --- | --- |
| Standard deviation | 4.3485 | 2.8538 | 2.4206 | 2.3905 | 1.8831 |
| Proportion of variance | 0.2055 | 0.0885 | 0.0637 | 0.0621 | 0.0386 |
| Cumulative proportion | 0.2055 | 0.2941 | 0.3578 | 0.4199 | 0.4584 |

**Supplementary Table 2.** Differentially expressed proteins between patients with influenza and patients with bacterial infections.

| **Protein** | **Influenza (n = 79)** | **Bacterial infections (n = 322)** | | **p-value** |
| --- | --- | --- | --- | --- |
| IL8 | 7.78536778481013 | | 8.5816851242236 | 0,000278 |
| CCL11 | 7.56094911392405 | | 7.20074456521739 | 3,98E-06 |
| CCL20 | 9.05340234177215 | | 10.5492861490683 | 3,00E-10 |
| CCL23 | 11.3601117721519 | | 12.0562967701863 | 9,43E-12 |
| CCL3 | 6.25293132911392 | | 7.49321649068323 | 4,34E-15 |
| CCL4 | 7.38342588607595 | | 8.48298155279503 | 3,45E-11 |
| CD40 | 12.5934606329114 | | 12.9362190993789 | 0,000358 |
| CXCL10 | 13.1371136075949 | | 11.7446202484472 | 1,90E-20 |
| CXCL11 | 11.73139 | | 10.2591934161491 | 5,45E-14 |
| FGF-23 | 4.46214069620253 | | 5.30152024844721 | 0,000442 |
| HGF | 10.4572824050633 | | 11.1182611490683 | 3,34E-07 |
| IFN-gamma | 10.5822778481013 | | 9.48106248447205 | 8,78E-05 |
| IL-17A | 2.94531563291139 | | 4.18545459627329 | 2,36E-11 |
| IL-24 | 2.24880151898734 | | 2.85692711180124 | 3,76E-06 |
| IL6 | 8.36291056962025 | | 10.0216559006211 | 6,97E-08 |
| LIF | 1.84252620253165 | | 3.12973860248447 | 4,59E-14 |
| MCP-2 | 11.808945443038 | | 10.2177758385093 | 8,99E-14 |
| MCP-4 | 14.4441601898734 | | 13.8177433850932 | 2,68E-05 |
| MMP-1 | 10.8696085443038 | | 11.4420218322981 | 0,000183 |
| OPG | 11.4179899367089 | | 11.7510125465839 | 0,000331 |
| OSM | 7.12268069620253 | | 7.94123782608696 | 1,90E-06 |
| SCF | 8.92490069620253 | | 8.53394726708074 | 0,000306 |
| TGF-alpha | 4.9039946835443 | | 5.88156913043478 | 3,80E-17 |
| TNF | 4.74096626582278 | | 5.51113894409938 | 5,58E-07 |
| TNFRSF9 | 7.34747436708861 | | 8.00897704968944 | 1,59E-06 |
| TNFSF14 | 5.73642911392405 | | 6.31825776397516 | 2,25E-08 |
| TRAIL | 8.1188232278481 | | 7.20759726708075 | 5,77E-15 |
| VEGFA | 6.80248393433742e-06 | | 11.7888084177215 | 6,80E-06 |

**Supplementary Table 3.** Differentially expressed proteins according to disease severity comparing patients with high SOFA score (>4) and patients with low SOFA score (<2).

| **Protein** | **SOFA score <2**  **n = 72** | **SOFA score >4**  **n = 62** | **p-value** |
| --- | --- | --- | --- |
| IL8 | 7.54511253424658 | 10.0506224193548 | 4.13823210801972e-13 |
| VEGFA | 11.9251419863014 | 12.6223388709677 | 1.49187362333609e-10 |
| MCP-3 | 3.38130623287671 | 4.45425661290323 | 3.66289033978312e-05 |
| CDCP1 | 4.51357815068493 | 4.98085096774194 | 0.000500836359188533 |
| OPG | 11.5039200684931 | 12.1871877419355 | 7.99483948196142e-09 |
| LAP TGF-beta-1 | 8.29018849315069 | 8.626335 | 0.000317647211401689 |
| uPA | 10.4486979452055 | 10.8321574193548 | 3.05162879708093e-05 |
| IL6 | 8.86928554794521 | 11.276035 | 1.39317027137116e-08 |
| IL-17C | 2.6196152739726 | 4.033145 | 2.6953516893144e-11 |
| MCP-1 | 13.2191141780822 | 14.1845708064516 | 1.00947687294954e-06 |
| IL-17A | 3.71904404109589 | 4.97488338709677 | 1.1639481020654e-05 |
| AXIN1 | 5.01794965753425 | 4.27246774193548 | 0.000735988023517331 |
| CST5 | 5.51110931506849 | 6.15005951612903 | 2.05813895879054e-05 |
| SLAMF1 | 2.16883678082192 | 2.61933064516129 | 4.1040070002183e-07 |
| TGF-alpha | 5.36495917808219 | 6.46573967741935 | 5.09686605947034e-09 |
| CCL11 | 7.10763821917808 | 7.43872774193548 | 0.000922599826720678 |
| FGF-23 | 4.19015582191781 | 7.05758241935484 | 7.75226544550476e-13 |
| FGF-5 | 1.52156767123288 | 1.80379822580645 | 2.80448223862097e-05 |
| LIF-R | 4.63568376712329 | 5.0726135483871 | 5.0757264192724e-07 |
| FGF-21 | 7.89396767123288 | 9.48193983870968 | 1.08627697613365e-05 |
| CCL19 | 10.7040767123288 | 11.7283843548387 | 9.12166070276976e-09 |
| IL-15RA | 1.96988150684932 | 2.79340741935484 | 6.43547608294061e-12 |
| IL-10RB | 6.44204938356164 | 6.8326964516129 | 1.09463172158329e-07 |
| IL-18R1 | 9.43435 | 9.98374274193548 | 1.47643179219557e-05 |
| PD-L1 | 7.83681493150685 | 8.63897822580645 | 4.19943850438217e-08 |
| CXCL5 | 11.300358630137 | 10.2860420967742 | 0.000125365074803325 |
| HGF | 10.849035890411 | 11.5256979032258 | 9.81866527526374e-05 |
| IL-24 | 2.2605798630137 | 3.85442806451613 | 1.69183896808857e-12 |
| MMP-10 | 9.26558061643836 | 10.0158332258065 | 2.44330240208854e-06 |
| IL10 | 7.55581910958904 | 9.54024225806452 | 5.42307099634175e-05 |
| TNF | 5.06843897260274 | 6.32501096774194 | 5.43325430470919e-05 |
| CCL23 | 11.8848928767123 | 12.3486670967742 | 4.04616129880677e-07 |
| CCL3 | 6.95585308219178 | 8.21465290322581 | 1.26654591461499e-05 |
| CD40 | 12.6932576712329 | 13.5788738709677 | 1.3824006239517e-11 |
| LIF | 2.40454287671233 | 4.36838129032258 | 3.03641088836534e-06 |
| CX3CL1 | 6.53158664383562 | 7.58355661290323 | 2.4191775014675e-13 |
| TNFRSF9 | 7.48065815068493 | 8.92591516129032 | 2.1438471275234e-09 |
| CCL20 | 9.63605212328767 | 11.3375485483871 | 1.24487781392583e-09 |

**Supplementary Table 4.** Differentially expressed proteins according to outcome comparing patients with worse outcome (in-hospital mortality and/or ICU admission) and patients with less severe outcome.

| **Protein** | **Less severe outcome (n = 307)** | **Severe outcome (n = 94)** | **p-value** |
| --- | --- | --- | --- |
| CCL19 | 10.9480729641694 | 11.4805571276596 | 1,92E-09 |
| CCL20 | 10.0244119543974 | 11.0063239893617 | 4,50E-07 |
| CCL23 | 11.8513747882736 | 12.1404715957447 | 0.000131 |
| CCL3 | 7.0633367752443 | 7.85481803191489 | 0.000216 |
| CD40 | 12.7870199022801 | 13.1354343617021 | 0.00043 |
| EN-RAGE | 3.94320885993485 | 4.41047367021277 | 0.000475 |
| FGF-21 | 8.31934986970684 | 9.19117138297872 | 0.0004255 |
| FGF-23 | 4.80573684039088 | 6.21529175531915 | 2,39E-08 |
| HGF | 10.85139 | 11.4343475531915 | 2,37E-08 |
| IL10 | 8.02952491856677 | 9.15281037234043 | 0.000536 |
| IL-17C | 2.96255576547231 | 3.53380026595745 | 9,85E-09 |
| IL-18R1 | 9.50566439739414 | 9.86210085106383 | 9,38E-09 |
| IL-24 | 2.53017377850163 | 3.41300531914894 | 0,00000791 |
| IL6 | 9.32308771986971 | 10.9090979255319 | 0,000000113 |
| IL8 | 8.10929723127036 | 9.45523845744681 | 0,000000261 |
| LIF | 2.5299790228013 | 4.00672170212766 | 1,79E-08 |
| MCP-3 | 3.56726726384365 | 4.33320771276596 | 2,37E-09 |
| OPG | 11.595607752443 | 11.978677287234 | 1,11E-09 |
| SCF | 8.73034951140065 | 8.22107313829787 | 2,39E-09 |
| TGF-alpha | 5.54186123778502 | 6.16946212765957 | 6,05E-08 |
| VEGFA | 11.9935900651466 | 12.3526069680851 | 1,79E-09 |
